# Efficient Citation Screening by Weak Classifier Ensemble*

**DOI:** 10.64898/2026.01.07.26343635

**Authors:** Xiaorui Jiang, Opeoluwa Akinseloyin, Vasile Palade

**Affiliations:** School of Information, Journalism and Communication, University of Sheffield, The Wave, 2 Whitham Road, Sheffield, S10 2AH; Centre for Computational Sciences & Mathematical Modelling, Coventry University, Puma Way, Coventry, CV1 2TT

**Keywords:** Automated systematic review, Citation screening, Large language model, Ensemble, Weakly supervised learning

## Abstract

Citation screening in systematic review is time-consuming. Machine learning can help semi-automate it but faces obstacles. Each systematic review is a new dataset without initial annotations. Extreme class imbalance against irrelevant studies makes it difficult to select a good subset of samples to train a classifier. The rigid requirement of a (near) total recall of relevant studies demands a careful trade-off between accuracy and recall. This paper pilots a weak classifier ensemble approach to tackle both challenges. The idea of ensembling is employed in two ways. First, multiple cost-effective large language models are applied and averaged to score and rank candidate studies to create a balanced pseudo-labelled training set. Second, different sets of pseudo-negative samples are bootstrapped from low-rank documents and multiple classifiers are trained and combined to make screening decisions. Experiments on 28 systematic reviews demonstrate significant performance improvements brought by the weakly supervised classifier ensemble, which also meets the rigid recall requirement for it to be safely used in practice.

## I. Introduction

Systematic review (SR) is the standard approach to build a comprehensive evidence synthesis of a research topic. One of the most tedious SR steps is citation screening—identifying all relevant studies that match the inclusion criteria of an SR. This step reduces the review size from several (dozens of) thousands to only a few (tens of) dozens [1]. There has been huge demand for using machine learning (ML) to (semi-)automate SRs and citation screening [2–3]. Citation screening is a very difficult ML task. A major challenge is caused by its cold-start nature— each SR about a different research topic constitutes a different dataset without initial inclusion/exclusion labels [4]. In both academic [5–7] and practical solutions [8–10], active learning (AL) has been the dominant technical paradigm. AL iteratively annotates samples and improves the screening performance of a classifier. The success of AL heavily relies on how fast it can find positive samples (i.e. included studies), which proves to be very difficult because most real-world SRs are extremely imbalanced towards excluded studies (i.e. negative samples). In addition, the requirement of a (near) total recall of all relevant studies, 95% at the minimum, is unbreakable in practice.

This paper pilots a novel lightweight approach for citation screening. The idea of ensembling is employed in two ways to tackle the aforementioned challenges. Firstly, the reasoning capabilities of large language models (LLM) in biomedical question answering [11] are employed to answer inclusion criteria questions, score and rank candidate studies following the ground-breaking work in [12], which provides a way to generate pseudo-labels for training a citation screener (Sect. II-A). To reduce costs and improve robustness, an ensemble of lightweight LLMs is used for this purpose, which proves to be effective. Secondly, the low-ranked side is used as a pool of negative samples. The idea of ensembling is employed by repeatedly sampling the negative pool to build multiple training datasets of pseudo labels, which are utilized to train multiple weak classifiers that are used in majority voting. This neat idea, called *weakly supervised classifier ensemble*, proves to be surprisingly cost-effective for building a citation screener that is capable of pre-screening a large portion of irrelevant studies, achieving significant workload reduction at a low cost.

This paper is organized as follows. Sect. II reviews the most relevant studies. Sect. III explains the proposed method step by step, including problem setup in Sect. III-A and the methods for scoring and ranking candidate studies using LLMs in Sect. III-B, for weakly supervised citation screening in Sect. III-C, and for building weakly supervised classifier ensemble in Sect. III-D. Sect. IV presents the experimental results and Sect. V concludes the paper with practical implications.

## II. Related Work

Machine learning for citation screening started with Cohen et al.’s seminal work [13], where versatile features are extracted to train a binary classifier and a 50/50 split of an SR dataset was used for evaluation. Later studies improved the screening performance by exploiting either more advanced features [14] or classification techniques such as deep learning [15]. These studies, however, require manual annotation of 50% of an SR dataset, which is unrealistic in real-world practice. Reducing the amount of manual annotation may not always work, as this may result in losing representative positive samples, causing significant damage to classifier performance.

Because citation screening is a cold-start problem, active learning has been extensively used in research [5–10] and most products of practical use. Active learning allows systematic reviewers to interact with the classifier by re-annotating some samples, either the most certain [5] or uncertain ones [6] by the classifier, and retraining the classifier until performance is satisfactory or saturated. However, this process is still tedious because first the initial classifier’s performance is poor due to lacking good methods for selecting a sufficient number of good positive samples; second, due to the above reason typically a large number of iterations are needed; and third, there is no reliable method to decide when to stop in order to guarantee satisfactory performance that meets the recall requirement.

The advent of LLMs, particularly ChatGPT in the first half of 2023, stimulated much hope for automating SRs, including citation screening, using LLMs’ impressive reasoning and question answering capabilities [16–18]. While it is argued that LLMs are not yet ready for automating SRs [19], they were proven to be strong screening prioritisation [12]. While an individual LLM, no matter how strong it is, cannot guarantee satisfactory performance, such as high accuracy and near-perfect recall for citation screening, combining multiple LLMs may significantly improve the robustness over individual LLMs [20–21]. Both lines of research inspired this paper.

## III. Methodology

### A. Problem Definition

A citation screening dataset for an SR is a collection of titles and abstracts of scientific articles, called *candidate studies*, defined as 𝒟 = {*d*_1_, ⋯, *d*_*N*_}. The SR protocol defines a set of inclusion criteria. Only the candidate studies that match all criteria will be *included* in the SR. Those failing to meet one or more criteria are *excluded* from the SR. Thus, the inclusion criteria can be converted into a set of YES/NO questions, denoted by *Q* = {*q*_1_, ⋯, *q*_*K*_}. The purpose of citation screening is to build a classifier that assigns each document a label *y* ∈ {0,1}, meaning “excluded” and “included”, respectively.

### B. Prioritisation by Lightweight LLM Ensemble

Given an LLM **M** as a question-answering engine, for each document *d* ∈ 𝒟, each inclusion criteria question *q* ∈ *Q* is answered by **M**, and the answer is formatted to begin with an *a*nswer keyword “Positive” (for an YES answer), “Negative” (NO), or “Unknown” (unable to answer or not having adequate information to answer), which is denoted by 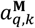, followed by a succinct explanation of the *r*eason for giving this answer, denoted by 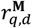. A question-level score for document *d* with respect to criterion *q*, denoted by *score*(*d, q*; **M**), is defined as the sentiment score calculated by BART [X], as in (1).

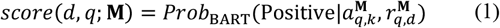

The document score with respect to inclusion criteria *Q* is defined as the average of all question-level scores as in (2).

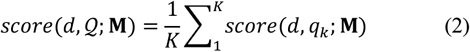

Reranking is introduced to account for the fact that information provided by included studies should be more relevant to what is asked in the inclusion criteria. For a document *d, question-level reranking* is done by averaging the question-level score and the semantic relevance between *d* and each criterion *q*, which is measured by the cosine similarity between the text embeddings of *d* and *q*, denoted by *rel*(*d, q*). *Document-level reranking* is done by averaging the document score and the semantic relevance between a candidate study d and the inclusion criteria paragraph *Q*, denoted by *rel*(*d, Q*).

𝒟 are ranked in descending order of reranking scores. To further increase ranking robustness, multiple LLMs are used to score each document, and the final score is the average of the scores of all LLMs. So, the LLM ensemble acts as a soft voter on individual LLM’s results. To maintain low financial and computational costs, this paper chooses a lightweight approach by combining multiple smaller and cheaper LLMs and will demonstrate the effectiveness of this approach.

### C. Weakly Supervised Citation Screening

Using the approach in Sect. III-B, the LLM ensemble scores each candidate study in 𝒟 based on how well they meet the inclusion criteria. This allows for creating a training dataset without the need for human annotation, which, to some extent, is akin to the idea of “LLM-as-a-Judge” [22]. Thanks to LLMs’ medical question answering capabilities, most included studies have higher chances of being ranked higher than excluded studies [4]. This paper employs the top *p*% of documents as “positive” samples pseudo-labelled by the LLM ensemble, denoted by 𝒯, and samples *b*% from the low-ranked documents as pseudo “negative” samples, denoted by ℬ, from the least ranked. A classifier is trained using the pseudo-labelled dataset 𝒯 ∪ ℬ. Although counterintuitive at first glance, such weakly supervised classifiers prove to be effective as a pre-screener.

If only the least-ranked documents are chosen to construct ℬ, which is a safe but conservative method, the accuracy of the resultant classifier is low, limiting the overall reduction of screening workload. Taking advantage of the large number of irrelevant studies (thanks to the extreme imbalance), a potential solution can be choosing low-ranked documents that are more distant from the bottom of the ranking list, which allows pushing the decision plane of the classifier closer to the top end. However, this will also increase the chance of excluding some relevant studies that are unfortunately ranked low by the LLM ensemble. Although such cases are rare, losing them may invalidate an SR especially if it has only few included studies.

A solution to kill two birds with one stone (i.e., increasing accuracy and precision while keeping perfect recall) is to build an ensemble of multiple such weak classifiers by sampling from a larger pool of potential negative samples, say the lower half of the ranking list, based on hypotheses that (1) there is a much higher chance of sampling truly irrelevant studies from the large pool (see [23] again for evidence) and (2) despite the fact that individual classifiers trained on such samples may ignore truly relevant studies that are ranked low, but a majority voting over a large number of diverse weak classifiers has a very good chance to correct most individual classifier mistakes. Based on these assumptions, this paper selects the least ranked *p*% (*p* > *b*) as the negative pool and repeatedly samples a small number of them to construct *L* versions of the set of pseudo negatives, denoted by 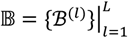. For each ℬ^(*l*)^, a classifier is trained on 𝒯 ∪ ℬ^(*l*)^. In total *L* weak classifiers are trained. Then, an ensemble is built over them using two strategies. *Soft voting* averages the posterior probabilities of *L* base classifiers for decision making and decides to include if the mean probability exceeds a threshold, 0.5 by default. *Hard voting* counts the number of inclusion decisions made by base classifiers and decides to include if the count exceeds a threshold, ⌊*L*/2⌋ by default. Ties are broken by mean probability.

## IV. Experimental Results

### A. Dataset and Metrics

Twenty-eight SRs in the test split of the 2019 Technology-Assisted Reviews in Empirical Medicine shared task [23]. Dataset details can be found in [12]. The total numbers of candidate studies to be screened are 39,792 for twenty SRs about clinical intervention (Int) and 26,830 for eight SRs about diagnostic technology assessment (DTA). Following [12], the inclusion criteria are converted into five questions for each SR. Experimental results are reported in two settings. In the *citation screening setting*, the weak classifier ensemble is used to make binary decisions and performance metrics are accuracy (Acc), precision (Prec), recall (Rec), f1-score (F1) and actual Workload Saving over Sampling (WSS)—the percentage of documents tagged as excluded. In the *screening prioritization setting*, the voter generates a new score to rank the documents and performance metrics are MAP (Mean Average Precision), WSS95 and WSS100 (WSS at 95% and 100% recall of include studies) [13], and R@*k*% (recall at top *k*%, *k* = 5, 10, 30, 50).

### B. Algorithmic Setup

Algorithmic options are chosen in a cost-effective manner. Three lightweight mainstream LLMs of moderate sizes are used for answering inclusion criteria: GPT-4o Mini (“gpt-4o-mini-2024-07-18”), Gemini 1.5 Flash (“gemini-1.5-flash-preview-0514”) and Claude 3 Haiku (“claude-3-haiku-20240307”). The temperatures are all set to 0 for reproducibility. For reranking, the GPT text embedding model “text-embedding-ada-002” is used. The classifier is linear support vector machine. Due to the limited space, the following parameter values from a broader set of experimental configurations are used for demonstrative purpose: *L* = 50, *t*% = *b*% = 2.5%, and *p*% = 10%.

### C. Results about Citation Screening

Table 1 shows the results in the citation screening setting. “w/o ensemble” is the single classifier trained with the top *p*% and bottom *b*% pseudo samples. Macro means are calculated by averaging the performance metrics of all SRs. Micro means are calculated by merging all SRs into one giant dataset. Soft voting significantly improves screening performance on all metrics. Notably, the relative improvement in macro mean accuracy is about 16.5% on Int (from 47.4% to 55.2%) and 15.5% on DTA (from 51.5% to 59.0%). Accordingly, macro mean WSS is increased from 41.8% to 49.7% on Int and from 44.3% to 52.0% on DTA, recording relative improvements of 18.9% and 17.4%. Micro means have a similar tendency, rising from 50.0% to 54.7% on Int (a relative 9.4% improvement) and from 44.3% to 55.2% on DTA (increased by 24.6%). Macro WSS, accordingly, is increased from 47.8% to 52.5% on Int and from 48.5% to 57.2% on DTA, recording 9.8% and 17.9% relative improvements, respectively. It is worth noting that the weak classifier ensemble by soft voting achieves the rigid 95% recall on all twenty-eight SRs, implying its *high potential for adoption by human reviewers in real-world practice*.

**TABLE I.** Performance Comparisons in the Citation Screening Setting (In Percentage)

| Category | Methods | Macro Mean [Min, Max] |  |  |  | WSS | Micro Mean |  |  |  |  |
| --- | --- | --- | --- | --- | --- | --- | --- | --- | --- | --- | --- |
|  |  | Acc | Prec | Rec | F1 |  | Acc | Prec | Rec | F1 | WSS |
| INT | w/o ensemble | 47.4 [17.6, 76.5] | 8.9 [0.6, 27.5] | 99.8 [98.5, 100] | 15.4 [1.1, 43.2] | 41.8 [7.3, 71.7] | 50.0 | 4.2 | 99.7 | 8.1 | 47.8 |
|  | w/ soft voting | 55.2 [24.0, 95.0] | 12.3 [0.6, 52.2] | 99.6 [97.0, 100] | 19.8 [1.3, 68.6] | 49.7 [15.4, 89.6] | 54.7 | 4.6 | 99.2 | 8.9 | 52.5 |
|  | w/ hard voting | 68.0 [33.3, 96.4] | 18.0 [0.8, 61.1] | 97.1 [78.8, 100] | 26.5 [1.6, 75.9] | 62.9 [21.7, 91.0] | 64.8 | 5.7 | 95.0 | 10.7 | 62.8 |
| DTA | w/o ensemble | 51.5 [36.2, 61.3] | 11.4 [0.5, 48.1] | 99.4 [97.4, 100] | 17.8 [0.9, 64.6] | 44.3 [26.9, 60.2] | 3.0 | 3.0 | 99.3 | 5.9 | 48.5 |
|  | w/ ensemble | 59.0 [40.9, 69.9] | 12.6 [0.5, 46.6] | 99.0 [95.3, 100] | 19.6 [1.1, 62.6] | 52.0 [26.1, 65.6] | 58.7 | 3.6 | 99.1 | 7.0 | 57.2 |
|  | w/ hard voting | 72.1 [61.5, 86.0] | 16.3 [0.7, 50.6] | 97.4 [89.5, 100] | 24.4 [1.4, 66.7] | 65.2 [30.3, 80.4] | 73.0 | 5.4 | 96.9 | 10.2 | 71.5 |

Comparatively, although hard voting achieves much higher accuracy and WSS, both macro and micro, it struggles to meet this unbreakable requirement for all SRs. There is one Int SR with a particularly low recall at only 78.8%, causing 14 (out of 66) relevant studies to be missed (SR ID “CD012551”). On DTA, the worst SR is “CD012233”, for which hard voting’s recall is 89.5%, missing 4 (out of 38) relevant studies. Figure 1 shows the performances of hard voting by varying the threshold. By increasing threshold, accuracy and WSS slightly decrease while recall increases. However, it is still difficult for hard voting to get a satisfactory recall although its WSS is much higher than soft voting. However, we argue that these results do not totally invalidate hard voting. Instead, it can be used for prioritisation and finding most relevant studies more quickly. After that, a good classifier can be trained or techniques in [24] can be applied to help detect the last few relevant studies.

**Fig. 1.**
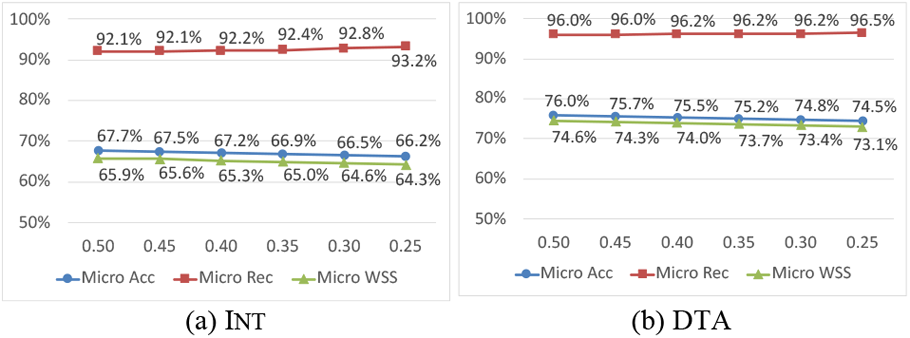
Impacts of threshold on hard voting performance.

It is vital to note that the proposed approach is designed to complement, not replace, active learning. It addresses several key challenges that hinder AL performance: (1) it provides a stronger initial classifier through a balanced pseudo-labelled training set, (2) it reduces severe class imbalance by efficiently filtering obviously irrelevant studies, and (3) it identifies more positive samples to initiate and accelerate the AL cycle.

### D. Results about Screening Prioritisation

It is interesting to see from Table 2 that both soft and hard voting’s performances are close to those of the LLM ensemble in ranking studies. Although slightly underperforming the LLM ensemble, they manage to train a much stronger classifier than the classifier that is trained using the LLM ensemble’s top and bottom ranked samples. These results demonstrate that on the one hand the weak classifier ensemble makes good sense because it does not distort the good results of the LLM ensemble and on the other hand it is the idea of ensembling that plays the key role in improving citation screening performance.

**TABLE II.** Performance Comparisons in the Screening Prioritisation Setting (In Percentage)

| Category | Methods | MAP | WSS95 | WSS100 | R@5% | R@10% | R@30% | R@50% |
| --- | --- | --- | --- | --- | --- | --- | --- | --- |
| INT | LLM ensemble | 47.0 [5.9, 91.4] | 69.6 [22.9, 94.4] | 63.6 [8.1, 99.4] | 56.6 [14.3, 100] | 71.8 [28.6, 100] | 91.5 [64.0, 100] | 97.9 [86.2, 100] |
|  | w/ soft voting | 42.2 [4.6, 94.9] | 68.8 [22.3, 92.8] | 65.0 [11.4, 97.8] | 54.9 [10.6, 100] | 71.5 [14.3, 100] | 89.4 [53.0, 100] | 97.3 [76.0, 100] |
|  | w/ hard voting | 44.1 [4.6, 96.1] | 68.0 [22.1, 93.1] | 64.7 [9.9, 98.1] | 57.8 [11.7, 100] | 73.7 [24.7, 100] | 90.0 [57.6, 100] | 96.3 [68.9, 100] |
| DTA | LLM ensemble | 35.9 [15.6, 58.3] | 70.8 [31.3, 92.3] | 66.3 [23.5, 97.3] | 56.5 [4.7, 100] | 72.3 [11.6, 100] | 88.6 [32.6, 100] | 96.6 [76.7, 100] |
|  | w/ soft voting | 32.0 [10.1, 57.3] | 68.6 [24.4, 89.6] | 63.2 [21.8, 94.6] | 54.1 [7.0, 90.9] | 68.5 [16.3, 100] | 88.0 [41.9, 100] | 96.4 [74.4, 100] |
|  | w/ hard voting | 34.0 [11.7, 60.4] | 70.6 [32.8, 90.1] | 64.7 [23.5, 95.1] | 55.6 [9.3, 100] | 69.5 [16.3, 100] | 89.3 [44.2, 100] | 97.0 [76.7, 100] |

## V. Conclusions

This paper introduces a novel approach for citation screening by employing a lightweight LLM ensemble for generating a pseudo-labelled dataset to train a citation screener. Ensembling is also employed to bootstrap a diverse set of pseudo negative samples for building a weak classifier ensemble. This neat idea has significantly improved the accuracy and workload savings in citation screening. With minimal hyperparameter tuning on twenty Intervention and eight DTA reviews, this approach achieves 52.5% and 57.2% screening workload reduction, respectively, with a near-total recall of relevant studies. It can be used as a cost-effective prescreening approach for systematic reviews and evidence synthesis. In addition, it alleviates class imbalance and finds positive faster, thereby better initializes and speeds up the active learning citation for citation screening.

## Data Availability

All data produced in the present work are contained in the manuscript

